# EFFECT OF FULL VACCINATION AND POST-COVID OLFACTORY DYSFUNCTION IN RECOVERED COVID-19 PATIENT. A RETROSPECTIVE LONGITUDINAL STUDY WITH PROPENSITY MATCHING

**DOI:** 10.1101/2022.01.10.22269007

**Authors:** Bumi Herman, Pramon Viwattanakulvanid, Azhar Dzulhadj, Aye Chan Oo, Karina Patricia, Sathirakorn Pongpanich

**Author notes:** **Registry**: NCT05060562. **Funder:** The Second Century Fund, Chulalongkorn University, Thailand.

## Abstract

**Background:** Symptoms after COVID-19 infection affect the quality of life of its survivor especially to the special senses including olfactory function. It is important to prevent the disability at an earlier stage. Vaccination as key prevention has been proven to be effective in reducing symptomatic disease and severity. However, the effects of vaccination on post COVID symptoms have not been evaluated. This study aimed to evaluate the possible protection of full vaccination and the occurrence of post-COVID olfactory dysfunction, specifically anosmia and hyposmia in patients who were diagnosed with COVID19.

**Method:** A longitudinal analysis using the retrospective cohort of the Indonesian patient-based Post-COVID survey collected from July 2021 until December 2021, involving COVID-19 Patients confirmed by RT-PCR and/or Antigen test. Variables including demography, comorbidities, health behavior, type of vaccine, symptoms, and treatment were collected through an online questionnaire based on the American Academy of Otolaryngology-Head and Neck Surgery (AAO-HNS). Participants were matched (1:1) using propensity matching score into two exposure statuses, infected 1)>14 days of full vaccination and 2)<14 days or incomplete or unvaccinated. The olfactory dysfunction was assessed two weeks and four weeks after negative conversion with PCR using a self-measured olfactory questionnaire (MOQ). The Generalized Estimating Equation (GEE) was performed to assess the effect of full vaccination on post-COVID olfactory dysfunction. The Receiver Operating Characteristic determined the sensitivity and specificity of the cutoff value of the days from fully vaccinated to diagnosis and the olfactory dysfunction.

**Results:** A total of 442 participants were extracted from the cohort and inoculated with the inactivated viral vaccine (99.5%). The prevalence of olfactory dysfunction in two weeks was 9.95% and 5.43% after four weeks. Adjusted by other variables, people who were infected >14 days after being fully vaccinated had a 69% (adjusted OR 0.31 95% CI 0.102-0.941) probability of developing olfactory dysfunction. Longer days of fully vaccinated to infection associated with increased risk (adjusted OR 1.012 95% CI 1.002-1.022 p-value 0.015). A cut-off of 88 days of full vaccination-to-diagnosis duration has Area Under Curve (AUC) of 0.693 (p=0.002), the sensitivity of 73.9%, and specificity of 63.3% in differentiating the olfactory dysfunction event in two weeks after COVID with a crude odds ratio of 4.852 (95% CI 1.831-12.855 p=0.001)

**Conclusion:** After 14 days of full vaccination, the protective effect could reduce the chance of post-COVID olfactory dysfunction although a longer full vaccination-to-diagnosis duration increases the risk. It is important to consider a booster shot starting from 89 days after the last dose in those who received the inactivated viral regimen.

## Introduction

### Epidemiology

The severe acute respiratory syndrome coronavirus 2 (SARS-CoV-2) infection causes Coronavirus Disease 19 (COVID-19) that has infected more than 270 million people as of mid-December 2021 with 5.3 million fatalities. In Asia, 44.7 million people were diagnosed with COVID-19 and 714,884 death cases were recorded. Indonesia leads the Southeast Asian region with 4.2 million cases and 143,960 mortalities (1). The COVID-19 manifestations also varied across individuals and severely affect people with comorbidities, particularly individuals with hypertension(2). The rapid transmission and high rate of mutation affect all aspects of life as the consequences of imposing stricter public health measures. The SARS-CoV-2 infection not only manifests through respiratory symptoms such as cough or fever(3) but also affects other organ systems, as well as alter the quality of life through the occurrence of persistent symptoms.

### Persistent covid symptoms definition and impact

The deterioration of quality of life can be seen in some COVID-19 patients as only 50-70% of the patient (either hospitalized or non hospitalized) are symptom-free after one month from disease onset(4, 5). It is important to define whether the collective persistent symptoms after covid should be treated as a syndrome or not. The term long COVID was defined by the National Institute for Health and Care Excellence (NICE), the Scottish Intercollegiate Guidelines Network, and the Royal College of General Practitioners as “signs and symptoms developed during or following a disease consistent with COVID-19 and which continue for more than four weeks but they are not explained by alternative diagnosis”(6)

An integrative classification shows that the timeframe of post-COVID symptoms is different between the hospitalized and non hospitalized patients. The transition phase in non-hospitalized patients is included since the onset of the symptoms, while among the hospitalized patients, the phase starts after hospital discharge. The transition phase may last up to 4-5 weeks. If symptoms continue longer than that, further classified into Phase 1:acute post-COVID (5-12 weeks), Phase 2: long post-COVID symptoms (12-24 weeks), and Phase 3: persistent post-COVID symptoms (more than 24 weeks)(7).

In a recent meta-analysis, olfactory dysfunction is listed as one of the post-COVID symptoms. Anosmia or loss of smell accounted for 21% of all cases observed for up to 3 months(8) A study involving a 6-month observation reported that the issues related to smell alteration is linked to self-confidence deterioration as people were unable to smell the fragrance, feel depressed, worry about body odor, and unable to enjoy the smell of food and worsen by the loss of taste. Around 43% of participants expressed depression. Therefore, preventing infection as well as handling the impact of olfactory dysfunction after COVID remain crucial(9). In Indonesia, a study involving a small number of hospitalized participants revealed that the loss of taste occurred among 56.8% of participants with a median of 3 days, and 68.9% of them were classified as severe smell disturbance(10).

### The pathophysiology of olfactory dysfunction in COVID

The nasal cavity consists of five cell types of the epithelial layer, in which the olfactory sensory neuron is responsible for detecting odorants(11). This cell creates synapses in the olfactory bulbs that connect to the axons and transmit the response into the olfactory cortex in the central nervous system (CNS). As the main entrance of SARS Cov2 into the body, the nasal cavity contains a higher level of Angiotensin-Converting Enzyme 2 (ACE2). The SARS-COV2 invades the cell by binding the viral spike protein to the human ACE2 receptor situated on the surface of the cell(12).

The olfactory dysfunction, such as hyposmia (partial loss of smell) and anosmia (total loss of smell) in COVID 19 patients occurs in two different mechanisms: the obstruction and the direct injury to the nasal cavity cells. The congestion is mainly caused by nasal epithelial edema which occurs temporarily. However, COVID-19 patients also reported anosmia without congestion. The olfactory epithelial injury is associated with pro-inflammatory cytokines as suggested by several studies including the alteration of TNF-α,IL-1β (13), or Interleukin 6 levels(14). Furthermore, emanating evidence have also reported olfactory nerve injury the injury of olfactory neuron although this cell does not express ACE2 receptors. The subsequent injuries of supporting cells play important roles in olfactory neuron injury(15). The occurrence of phantosmia (distorted sense of smell) and olfactory hallucination (perceived distortion in the absence of an odorant) in COVID-19 patients suggest the involvement of the CNS, either the virus enters through the olfactory bulb or disseminates in the bloodstream invading the endothelial blood-brain barrier(16)

### Vaccination situation in Indonesia

A number of suggested therapies for olfactory dysfunction have been proposed by many studies including olfactory training and steroid regimen and have shown benefits in Postviral olfactory dysfunction(17). However, it is important to note that the prevention of COVID-19 by vaccination is by far the most appropriate way to reduce the burden of post-COVID-19 olfactory dysfunction. In Indonesia, by December 10^th,^ 2021, 53% of the targeted population received at least one dose of vaccination while only 37.2% received two doses(18). The primary regimen In Indonesia is two doses inactivated vaccine 14-28 days apart, and viral vector vaccine with 8-12 weeks apart. The mRNA vaccine recently given as a booster to medical personnel in the middle of 2021 and for those who are unable to receive the primary vaccine. This proportion is still lower than expected.In addition, the waning protection of full vaccination is a new threat where the need for a booster for the common population is now being considered.

### Objective and Hypothesis

It is important to assess the subsequent effect of vaccination in preventing olfactory dysfunction as one of the post COVID-19 symptoms. Furthermore, this study could enhance the need for accelerating vaccination programs or considering booster shots and the ideal time to provide the booster shot. The initial assumption is, despite being infected by SARS COV2, people who have been fully inoculated have a lower probability of developing post-COVID-19 olfactory symptoms.

## Methodology

### Setting and Study Design

This study utilized the Indonesian POST-COVID retrospective longitudinal data, involving participants from the entire provinces of Indonesia, collected until December 2021. Participants were requested to fill the questionnaire which wasdelivered through an online invitation link. The source of information was collected from the telemedicine records and daily observation documented by the individual. Additional clinical information in the form of a medical resume was also obtained if the participants underwent hospitalization.

### Participants Eligibility

This study recruited participants at any age as long as the subject was able to fill and answer the questions provided. The questionnaire was disseminated to groups of COVID 19 survivors and the snowball technique was conducted to obtain more participants. Furthermore, dissemination through social media was performed by the covidsurvivor.id, a large COVID-19 survivor group in Indonesia. All participants must be diagnosed as COVID 19 using the Real-Time Polymerase Chain Reaction/PCR (ref), other Nucleic Acid Amplification Test/NAAT(19), or the rapid antigen test (20)of the nasopharyngeal sample as the sample from this area shows higher sensitivity and specificity for SARS COV2 detection(21). Moreover, the participants should be declared as cured for at least two weeks by the physician. The definition of cured in COVID-19 is based on the World Health Organization criteria for releasing COVID-19 patients from isolation treatment(22). However, this standard is unable to capture the conversion of PCR results particularly for those who were not hospitalized. Therefore, in this study, participants with negative PCR results and suggestive clinical recovery are considered cured. Participants were excluded if they had missing repeated outcomes and a history of suspected reinfection.

### Variables

The authors classified the variables into three groups. Firstly, the demographic factors consist of age at diagnosis, sex at birth, occupation (medical staff or not), education, and province of domicile. Secondly, the general health status including Body Mass Index (BMI, calculated as body weight in kilogram divided by the square of body height in meter) during COVID 19 episode, presence of chronic disease and comorbidities, smoking, alcohol drinking within three months before diagnosis, and moderate physical activity (which defined as 30-40 minutes of physical activity involving a warm-up, main session, and cool-down) were also obtained. The third group of variables is related to the first COVID 19 episode including the time of diagnosis, and method of diagnosis confirmation. Participants were asked about the symptoms and duration of the symptoms, medication given, any oxygen supplementation required, whether the participant was hospitalized, or received intensive care or receiving plasma convalescent therapy in their COVID 19 episode.

### Tools

The general questionnaire used by the main cohort is linear with the COVID 19 Anosmia Reporting Tool developed by the American Academy of Otolaryngology-Head and Neck Surgery (AAO-HNS) with detailed information in treatment, the status of comorbidities, and time of onset. The COVID 19 diagnosis was based on the Real-Time (Polymerase Chain Reaction) PCR of gene ORF, or N, or Spike which is responsible for the SARS-COV2 infectivity(23) or antigen tests for those with symptoms or at higher risk (close contact) following the WHO recommendation.

### Exposure

This study defined the exposed group as **fully vaccinated and infected more than 14 days** after fully vaccinated, as, after this period, full vaccination is considered to have a protective effect from SARS-COV2 infection of more than 80%(24). Those people **infected less than 14 days after full vaccination or who did not receive full vaccination (either imcomplete or unvaccinated)** were considered as a non-exposed group. Type of vaccine was asked from the participants.

### Outcomes

The authors assessed the olfactory dysfunction according to the following duration; two weeks and four weeks after being declared cured to accommodate the acute post COVID period; and measured using the Self-Mini Olfactory Questionnaire (Self-MOQ). This 14-question tool is reliable to screen the olfactory dysfunction compared to the objective psychophysical olfactory examination(25) with the total score ranging from 0-14. The outcomes were coded into binary response following the cutoff score of 3.5 (25) to distinguish the normosmia (no olfactory dysfunction) from anosmia and/or hyposmia (olfactory dysfunction).

### Study Size and Possible Bias

To extract the eligible participants from the cohort and reduce selection bias, the authors applied a propensity matching score to match the participants into two groups. A 1:1 ratio was set, and variables to be included in the propensity analysis were occupation, education, island, type of living area, living companion, age, and hypertension status. These variables were selected to ensure the representativeness of the subset and the probability of getting the vaccination. Exact and fuzzy matching with a match tolerance value of 0.25 was set to match exposed and non-exposed participants. This yielded a bigger participant number compared to stricter match tolerance values (0.05 and 0.1).

The sample size is calculated considering 5% type I error, 95% power, with equal sample size between two groups and two repeated measures, assuming the correlation between measurements is 0.5. Effect size Cohen d was estimated, assuming that the probability of olfactory dysfunction is 25% higher among people who were not fully vaccinated or fully vaccinated less than 14 days. This led to a total sample size of at least 392.

. At the beginning of the questionnaire, the authors informed the participants to rely on their recorded observation to reduce the possibility of recall bias. Having a COVID 19 was an unforgettable experience for the participants, so they might recognize their symptoms better compared to having seasonal flu. Furthermore, the presence of telemedicine during home isolation reduced the recall bias since the participants were expected to record the signs and symptoms daily. The authors acknowledged that the symptoms that appeared after COVID 19 may or may not be related to COVID 19, hence, the participants were expected to consult their symptoms to the physician to ensure the reliability of their responses.

### Quantitative Variable

Several quantitive data will be discretized accordingly and following the common classification such as Body Mass Index. Participants who have never smoke are defined as Brinkman Index value 0, and more than 0 as a smoker. Days from the second shot to the diagnosis was also obtained by subtracting the date of receiving the second dose from the date of diagnosis.

### Statistical Analysis

Data cleaning was conducted without imputation as only complete responses would be received. The authors performed descriptive statistics and normality tests to determine the distribution of quantitative data. A bivariate analysis was made between each independent variable and the exposure status to ensure the similar characteristic between the group.

A Generalized Estimating Equation was selected to analyze the effect of full vaccination on the olfactory dysfunction after COVID. A repeated measure of two weeks and four weeks after being cured was set as the outcome. The covariance matrix followed the robust estimator. The binary response with the logit link function was applied. The Quasi Likelihood Under Independence Model Criterion (QIC) was used to select the best correlation matrix structure based on the lowest value, computed using the full log quasi-likelihood function. Several variables were included for adjustment following the bivariate analysis and collinearity assessment. Type III test of effect was conducted based on Wald Chi-Square. The confidence interval of the odds ratio was set at 95%.

The average time from full vaccination to infection is calculated and set as the cutoff. A discretization was made to separate the cohort into two groups based on the cutoff. A Receiver Operating Characteristic curve was conducted to determine the sensitivity and specificity of this cutoff in discriminating the olfactory dysfunction among fully-vaccinated people. A crosstabulation was executed between the discretized variable and the olfactory dysfunction event to obtain the crude odd ratio.

### Ethical Approval

This study was approved by The Research Ethics Review Committee for Research Involving Human Research Participants, Faculty of Medicine Hasanuddin University (Approval number of full review UH21110687). The author ensured that the data remain unidentifiable to protect the confidentiality of the participants and was used accordingly. This research is registered in clinicaltrials.gov number NCT05060562.

## Results

### Baseline Characteristic

A total of 442 participants were extracted from the cohort and equally allocated into the exposed and non-exposed groups. 220 (49.8%) people were diagnosed using PCR test, 117 (26.5%) with antigen test, and the rest with a combination of multiple tests. Since the data was taken retrospectively, the status of vaccination at the time of entering the cohort was different compared to the time of diagnosis.. The selection of participants is illustrated in figure 1. The proportion of alteration of smell two and four weeks after cured was 9.95% (44 cases) and 5.43% (24 cases), respectively.

**Figure 1.**
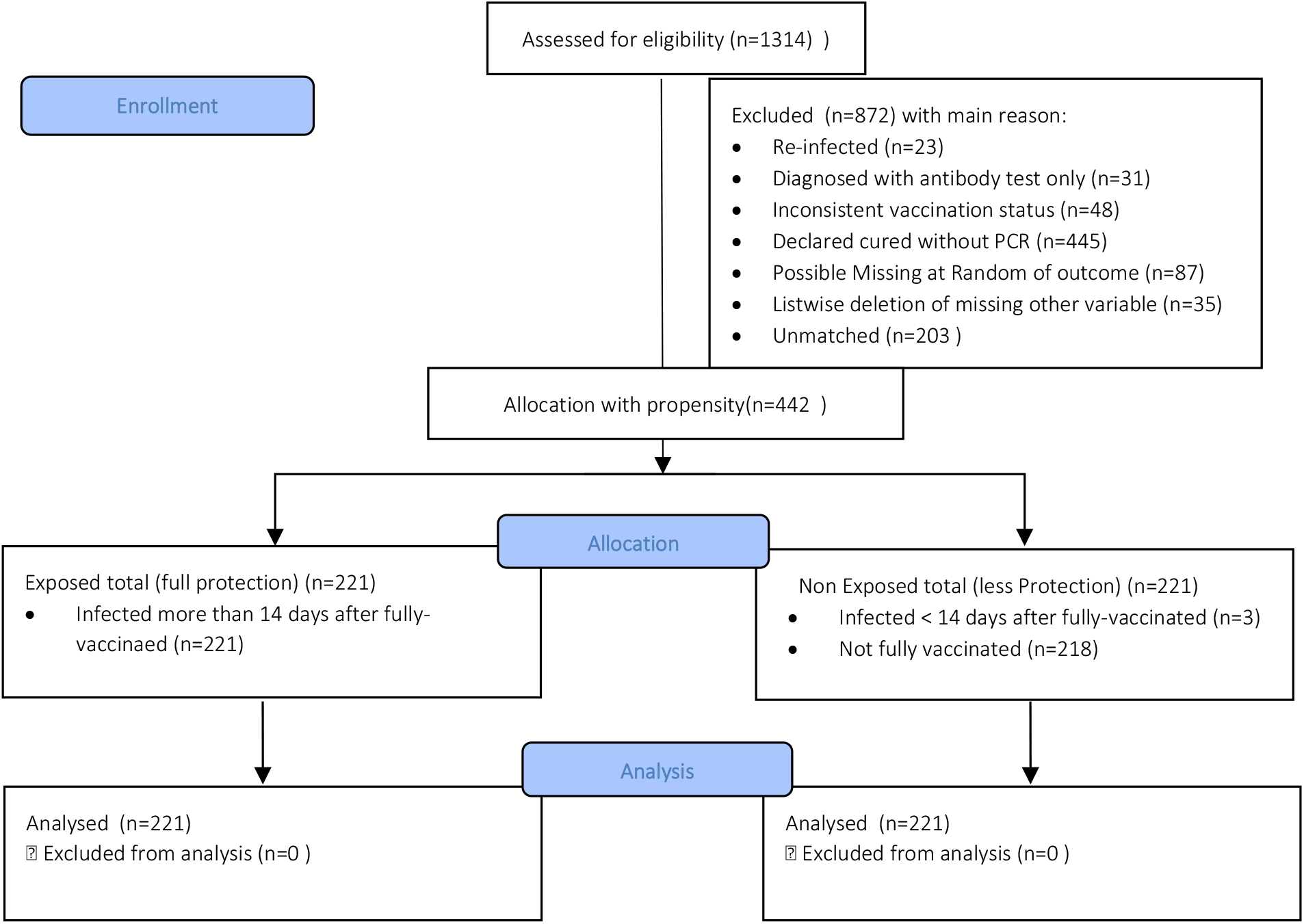
The participants’ flowchart

**Figure 2.**
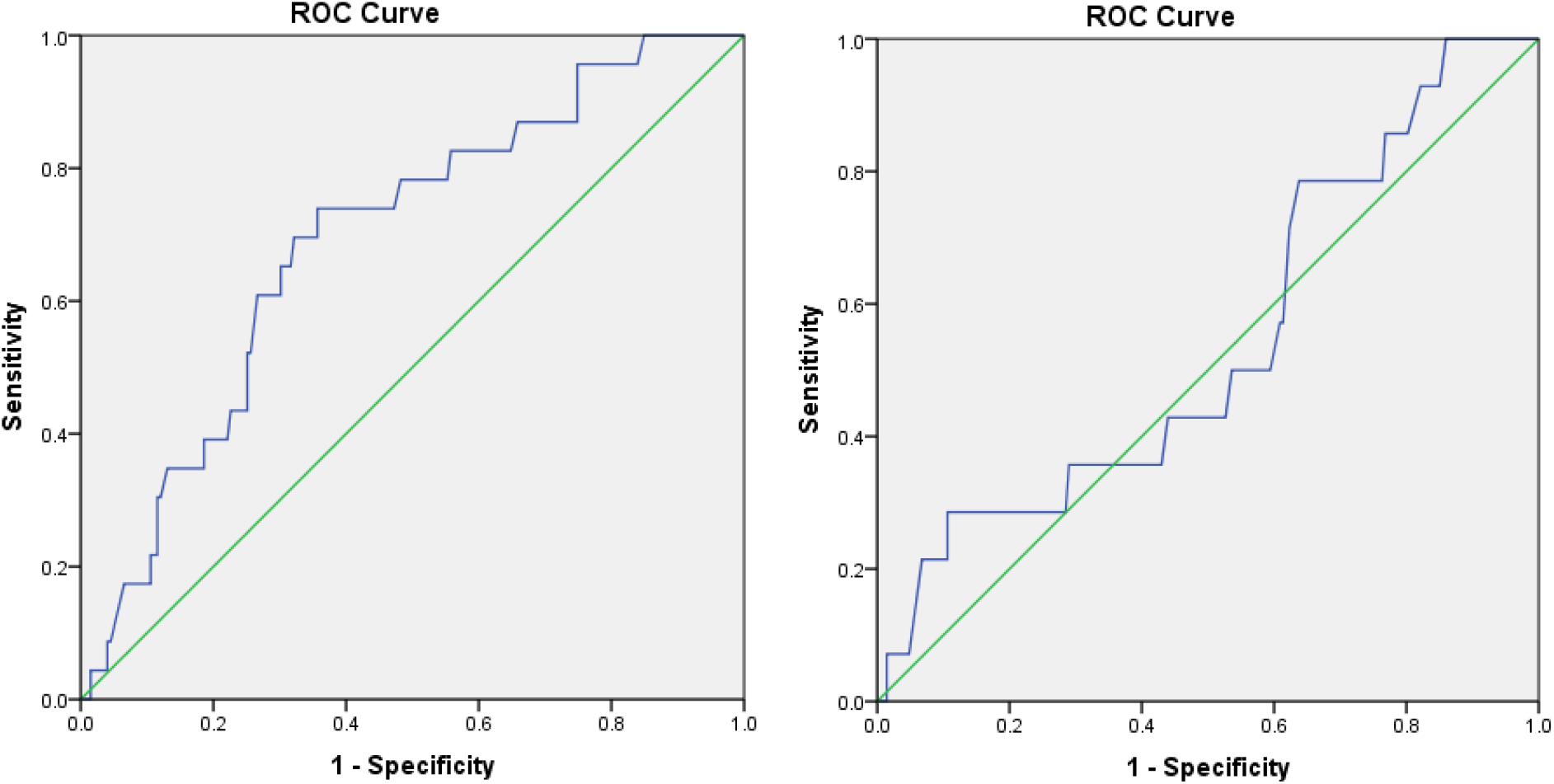
The ROC value for number of days from fully vaccinated to infection. The left figure demonstrates the Area Under Curve for the olfactory dysfunction event in two weeks after COVID, and the right figure for the olfactory dysfunction event in four weeks after COVID

The vast majority type of vaccine for inoculation was the 2 doses inactivated viral vaccine in all participants in the exposed group with only 1 participant fully inoculated with the viral-vector vaccine in 56 days. The average period to reach full vaccination in this group with the inactivated viral vaccine was 24 ± 6.41 days.

The baseline characteristic of the participants in Table 1 shows the different characteristics in terms of occupation where medical personnel mostly in the cohort (4% 240/442). The distribution of cases was dominant in Java island (35.9% 159/442) as it is the most populated island and where most of the cases were detected. The majority of cases were found in the capital of the province. Participants mostly stayed with other people before getting infected (68.1% 301/442).

**Table 1.**
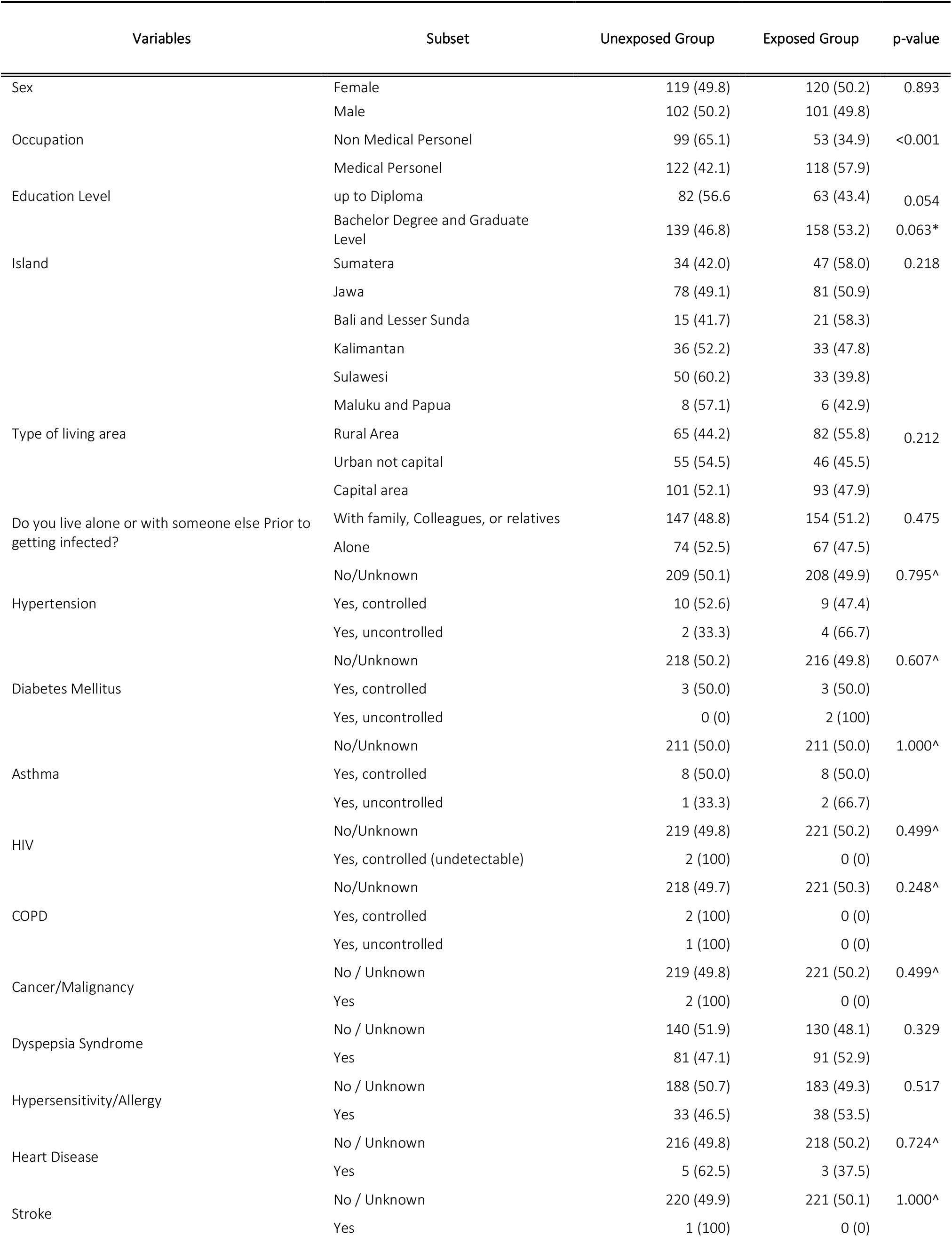
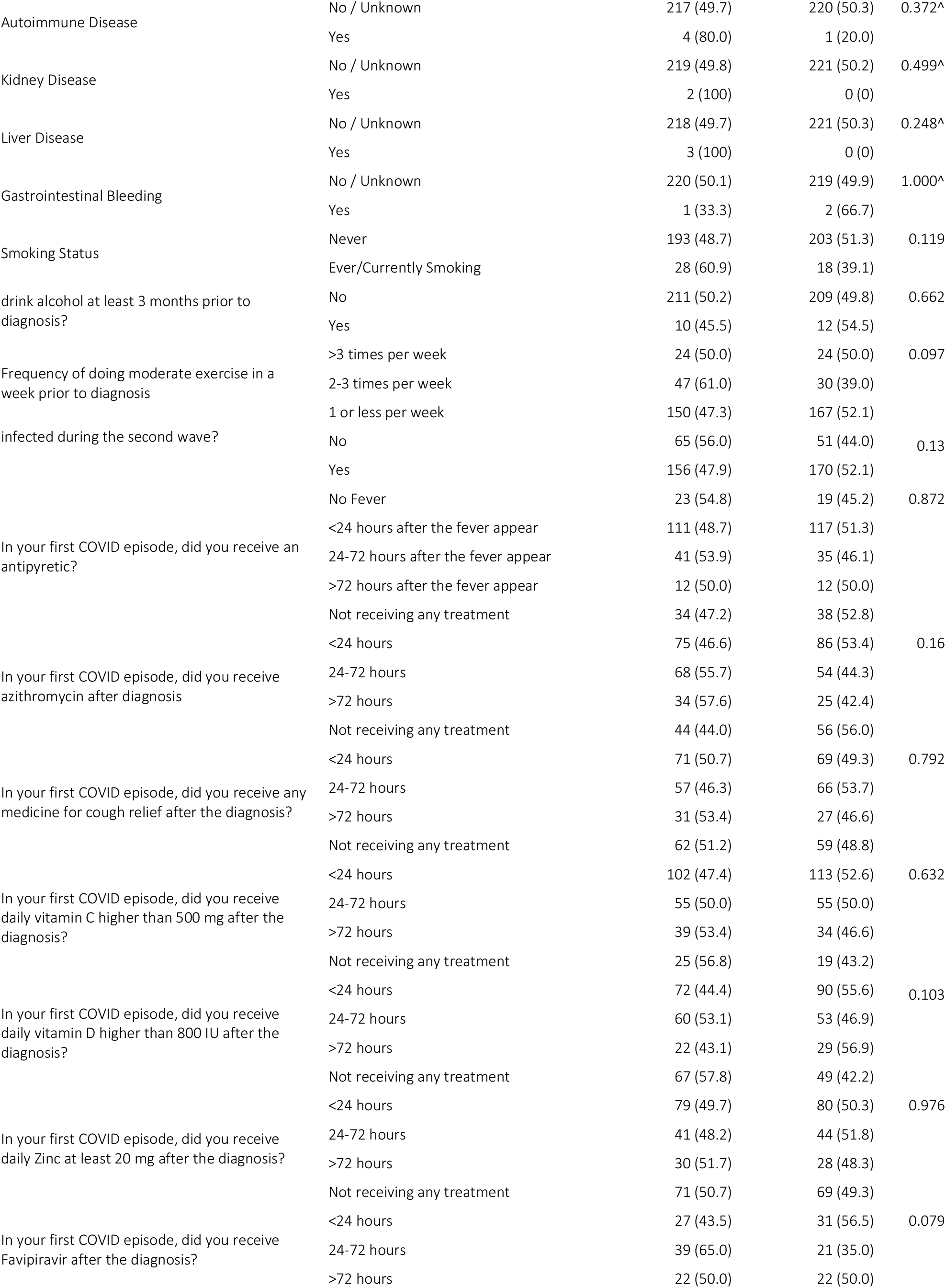
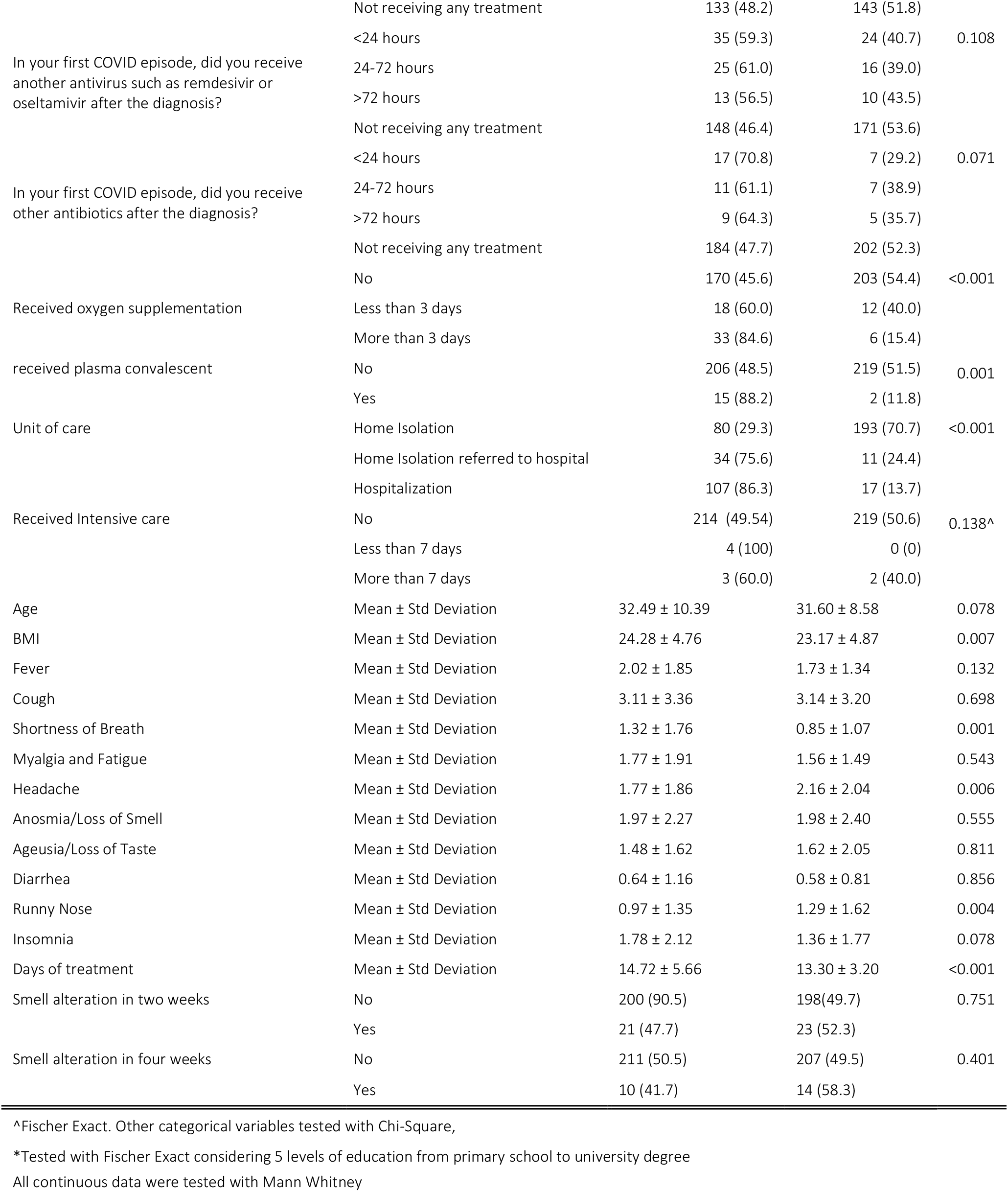
Baseline Characteristic

A total of 14 diseases and symptoms were asked and categorized by the level of current status. There was no difference between the presence of these diseases with the exposure status (p>0.05). In terms of health behavior, smoking, drinking alcohol, and moderate physical activity were similar across groups. However, the mean BMI was significantly different (p<0.007). The second wave of COVID 19, driven by the Delta variant occurred within 1 June – 30 September 2021 and most of the participants were infected during the second wave. There was no difference in the proportion of cases according to the COVID wave between exposure status(p>0.05).

In terms of total days of symptoms, there was a significant difference in shortness of breath (p=0.001), headache (p=0.006), runny nose (p=0.004), and days of treatment(p<0.001). Shortness of breath (1.32 ± 1.76 versus 0.85 ± 1.07, p=0.001) and days of treatment (14.72 ± 5.66 versus 13.30 ± 3.20, p<0.001), were shorter in the exposed group, although runny nose and headache were longer in the exposed group. Loss of smell and loss of taste duration were similar between groups (p>0.05). The average day of treatment was around 14 days, hence the two-week and four-week measurement would accommodate the timeframe of long COVID or transition phase, and acute post-COVID phase in integrative classification.

Standard COVID medication initiation during the first COVID-19 episode was identified according to the time of confirmed diagnosis and the time of onset. No significant difference was seen between the two groups (p>0.05). There was a significant difference in unit of care and advanced treatment (oxygen supplementation, plasma convalescent, and intensive care) between-group (p<0.001).

As the author assumed that there would be collinearity between the symptoms and the treatment, the duration of shortness of breath was selected as the main predictor to represent other significant parameters. Table 2 demonstrates the association between the duration of shortness of breath with other variables.

**Table 2.** Correlation between shortness of breath and other predictors

|  | Variables | Spearman rho | p-value |
| --- | --- | --- | --- |
| Shortness of Breath | Headache | 0.492 | <0.001 |
|  | Days of treatment | 0.488 | <0.001 |
|  | Body Mass Index | 0.079 | 0.019* |
|  | Runny Nose | 0.306 | <0.001 |
|  | Oxygen Supplementation |  | <0.001^ |
|  | Unit of Care |  | <0.001^ |
|  | Received Azythromicin |  | <0.001^ |
|  | Received Plasma Convalescent |  | <0.001& |

All significant at the 0.01 level (2-tailed) except \*
^Kruskal Wallis
&Mann Whitney

Following the baseline analysis and omitting factors with higher collinearity, the GEE analysis was conducted. Parameters included in the model were exposure status, repeated measurement, and interaction between the measurement and exposure status. Occupation and days of shortness of breath were treated as other factors. The authors also included days of getting infected since fully vaccinated, the island, and infected during the second wave as an additional parameter. Table 3 shows the parameter estimates.

**Table 3.** Parameter Estimates of The Model

| Parameter | B | Std. Error | Hypothesis Test |  |  | Exp(B) | 95% Wald Confidence Interval for Exp(B) |  |
| --- | --- | --- | --- | --- | --- | --- | --- | --- |
|  |  |  | Wald Chi-Square | df | Sig. |  | Lower | Upper |
| (Intercept) | -1.678 | 0.53 | 10.02 | 1 | 0.002 | 0.187 | 0.066 | 0.528 |
| Infected >14 days after fully vaccinated | -1.172 | 0.5672 | 4.271 | 1 | 0.039 | 0.31 | 0.102 | 0.941 |
| Week 4 after cured (reference week 2) | -0.808 | 0.3396 | 5.655 | 1 | 0.017 | 0.446 | 0.229 | 0.868 |
| Days from fully-vaccinated to infection | 0.012 | 0.005 | 5.863 | 1 | 0.015 | 1.012 | 1.002 | 1.022 |
| [Exposed] * [assessment 4<sup>th</sup> week] | 0.248 | 0.4536 | 0.298 | 1 | 0.585 | 1.281 | 0.527 | 3.116 |
| [Exposed] * [assessment 2<sup>nd</sup> week] | 0<sup>a</sup> | . | . | . | . | 1 | . | . |
| [Unexposed] * [assessment 4<sup>th</sup> week] | 0<sup>a</sup> | . | . | . | . | 1 | . | . |
| [Unexposed] * [assessment 2<sup>nd</sup> week] | 0<sup>a</sup> | . | . | . | . | 1 | . | . |
| Total days of shortness of breath | -0.217 | 0.1396 | 2.426 | 1 | 0.119 | 0.805 | 0.612 | 1.058 |
| Medical Personnel | -0.331 | 0.3446 | 0.925 | 1 | 0.336 | 0.718 | 0.365 | 1.411 |
| [Island = Maluku and Papua] | -0.082 | 0.6893 | 0.014 | 1 | 0.905 | 0.921 | 0.239 | 3.558 |
| [Island= Sulawesi] | -1.01 | 0.5292 | 3.64 | 1 | 0.056 | 0.364 | 0.129 | 1.028 |
| [Island=Kalimantan] | -0.98 | 0.5189 | 3.566 | 1 | 0.059 | 0.375 | 0.136 | 1.038 |
| [Island= Bali and Nusa Tenggara] | -0.561 | 0.5525 | 1.029 | 1 | 0.31 | 0.571 | 0.193 | 1.686 |
| [Island= Jawa] | -0.42 | 0.4045 | 1.079 | 1 | 0.299 | 0.657 | 0.297 | 1.452 |
| [Island=Sumatera (reference)] | 0<sup>a</sup> | . | . | . | . | 1 | . | . |
| Infected during second wave | 0.506 | 0.3917 | 1.666 | 1 | 0.197 | 1.658 | 0.769 | 3.572 |
| (Scale) | 1 |  |  |  |  |  |  |  |

For people who still get infected after being fully inoculated for more than 14 days, the chance of developing olfactory dysfunction was lower to 69% (adjusted OR 0.31 95% CI 0.102-0.941), although there was no significant interaction between the full vaccination status and the occurrence of symptoms in two and four weeks after recovery (p=0.585). Interestingly, the longer the days from fully vaccinated to infection, the higher the chance to develop the symptoms (adjusted OR 1.012 95% CI 1.002-1.022 p-value 0.015).

Following the significant effect of the number of days from fully vaccinated to infection, the authors conducted further analysis. The average vaccine-to-infection time in the exposed group was 88.36 ± 42.88 days. The Receiver Operating Characteristic curve of the nearest value from 88.36 (88.5) value shows the Area Under Curve (AUC) of 0.693 (p=0.002), the sensitivity of 73.9%, and specificity of 63.3% in differentiating the olfactory dysfunction event in two weeks after COVID. The specificity slightly changed after excluding one non-inactivated virus vaccine recipient (63.5%). However, insignificant results were seen when differentiating the event after four weeks (p>0.05)

Participants were then split into two groups,1) those who were infected more than 88 days after full vaccination and 2) less than 88 days and not fully vaccinated (the days considered as 0). The chi-square test shows an association with the olfactory dysfunction after two weeks (Crude Odds Ratio 2.803 95% CI 1.452-5.412 p=0.002). The Cochran Mantel-Haenszel test was conducted to see the odds ratio across the exposure status. Among the exposed group, the odds ratio inflated to 4.852 (95% CI 1.831-12.855 p=0.001). Contrary to the first results, no significant association with the olfactory dysfunction after four weeks of COVID resolution.

## Discussion

### Cohort Selection and Representativeness

This study addressed the effects of full vaccination on the occurrence of acute-post COVID-19 olfactory symptoms in Indonesia with an assumption that vaccination will reduce the infection and hence, the symptoms. The data represents the true proportion of cases in Indonesia (26), including the type of treatment where the vast majority of the cases were treated at home (273/442 61.8%), and also included several cases that occurred in 2020. Education level was almost significant, indicating that this online questionnaire might only reach highly-educated people and other people with lower education could come from the family or colleagues. The underlying diseases and the treatments prescribed were similar in both groups after matching. The prevalence of olfactory dysfunction accounted for almost 10% in two weeks but was reduced by half after four weeks. This finding is lower than the global estimation(8), however, the shorter duration of anosmia of lower than two weeks in an Indonesian study(10) supports the finding of why only 10% of participants experienced olfactory dysfunction after two weeks. In this study, people who are fully vaccinated and infected more than 14 days after that, have a lower chance to develop olfactory dysfunction which is linear with the prior assumption.

Propensity matching score was preferred over other techniques to adjust the potential confounder. However, some factors persist as potential confounders. In this subset, the proportion of medical personnel was higher than the true situation. One of the underlying reasons was medical personnel are the most susceptible group (27) and prioritized for full vaccination, therefore it is difficult to find a similar proportion to the true setting after matching. Propensity matching by hypertension is important as the blood pressure assessment is mandatory in the vaccination center and less invasive, therefore the information is more reliable. Furthermore, hypertension is associated with a higher risk of contracting COVID 19.

To select more participants. the initial plan was to do a case-control matching with unequal samples such as 1:2, however, stricter eligibility (particularly the requirements to do post-care PCR to eliminate the heterogeneous definition of cured) leads to lower available cases for such allocation. Post-care PCR is not required for those with mild cases and can be released form isolation after several days. In a real setting, post-Care PCR is conducted for non-medical purposes including work, or travel, or the patient fell into the high-risk group of getting the severe disease, and those who were hospitalized. A trade-off between a clear-cut definition of cured and the sample size explained the significant reduction of recruited participants from the general cohort.

In terms of vaccine type, the participants in the exposed group were inoculated with inactivated viral vaccine and only one participant received an adenovirus-viral vector vaccine. This is could reduce the generalizability of the finding, to only those who received inactivated viral vaccines. However, five people were infected after receiving the first dose of viral vector vaccine with the minimum days of 38 and the average of 46 days, thus indicating a long period of inoculation is a higher risk of being exposed to SARS Cov2. One dose of vaccine could have 56% protection from SARS Cov2 infection(24). One of the problems in recruiting people with a different type of vaccine is the procurement of vaccine in Indonesia, where the viral-vector vaccine and mRNA vaccine were not widely available for people at the period of recruitment. The sudden decrease of the case in Indonesia also hinders the recruitment of people who are inoculated by different vaccines and get infected.

### Quality of Information and Subjective Assessment of Outcome

The hospital-based study would not address the true effect of vaccination as those people who were vaccinated will experience milder symptoms and this is why the authors did the patient-based study. However, the retrospective data collection from the patient is not free from bias despite the researchers expecting the participants to rely on their written daily observation or the telemedicine record (which integrated into COVID service in 2021). Furthermore, olfactory dysfunction is difficult to assess objectively and clinical confirmation (which requires people to go to the hospital) is not feasible during the pandemic. This is the reason why this study conducted a self-assessment for olfactory dysfunction, which addressed as a limitation. The Quick Olfactory Sniffin’s Stick test is one of the objective assessments to confirm the olfactory dysfunction using certain strong scents put at the distance of 1-2 cm from nostrils and it has a good performance in identifying the olfactory dysfunction in COVID 19 patients(28). But the Self-MOQ underwent validation through Sniffin’s stick test and shows a good discriminant ability(25). Missing at random in outcome assessment was likely occurred as the entire questionnaire is long and the proposed analysis using GEE will introduce bias if the cases with missing data are included.

### Analysis and Effect of Vaccination

Aside from occupation, this study also considers island as important demographic factors due to distribution and procurement of vaccine which affect the allocation of exposure status, and the distribution of dominant SARS-Cov2 strains wherein Java during the second wave, Delta variant was the most prevalent strain, and small percentage Alpha and Beta were found in Sumatera and Java(29). Mutation of SARS-Cov2 leads to different clinical manifestations(30) due to different abilities to attach to the host cell or the ability to escape from the immune response. Furthermore, Indonesia experience two COVID-19 waves where the first wave was dominated by wild type variant followed by Alpha and Beta. Several studies indicate the decremental vaccine efficacy against each variant(31). Indonesia deployed inactivated vaccine as the primary regimen for vaccination, which is based on wild type variant, therefore, people who were infected in the second wave may have detrimental protection. This was then included as further adjustment.

The treatment given to the patient was according to the guideline, where the main antivirus was Oseltamivir, particularly for mild cases. Favipiravir was available later during the second wave and remdesivir is mainly administered in the hospital. Vitamin and mineral supplementation is given as a package in telemedicine or field hospitals. Currently no clear association or no supporting study between administration of Zinc(32), Vitamin C, Vitamin D, or even plasma convalescent and subsequent olfactory dysfunction. In this study, there were no associations between all of these treatments and the olfactory dysfunction within two weeks(p>0.05), except Azithromycin administration <24 hours after diagnosis and olfactory dysfunction in two weeks using a simple logistic regression model (OR 0.311 compared to not receiving therapy at all 95% CI 0.132 0.734 p value=0.008). Aside from antibacterial properties, Azithromycin also has antiinflammation properties through a dose-dependent lymphocyte activity and proinfllmataory cytokines secretion(33) However, there is a strong association between azithromycin adminsitration and days of shortness of breath (Table 2). Azithromycin used to be administered in all cases, but the updated guideline suggests azithromycin when there is a clear indication including pneumonia.

The duration of shortness of breath represented other factors based on the collinearity test, therefore in the final analysis, the shortness of breath acted as the factor that reflects the severity of disease(34), assuming that severe disease will also lead to sequelae and persistent symptoms.

Calculation of the crude odds ratio of vaccination and olfactory dysfunction from table 1 reveals an interesting point that people who were fully vaccinated more than 14 days were actually had a higher risk of developing olfactory dysfunction in two weeks (OR 1.11) and four weeks (OR 1.43) from cured. This is why several factors should be taken into account for final analysis. The final model reveals the protective effect of full vaccination against olfactory dysfunction. However, it is important to notice that the waning of vaccine protection in people who were infected more than 88 days after being fully vaccinated leads to a higher chance of developing olfactory dysfunction than people who infected less than 88 days.

### Strength and Limitation

The author applied a robust method to adjust the flaw of the retrospective design and potential confounders. The propensity matching score reduced the selection bias. Rigorous discretization of variables and the robust analysis using GEE estimates the effect in an unbiased way. However, clinical confirmation of the information provided by participants is crucial, such as obtaining the true value of blood glucose to determine the status of diabetes and define other chronic diseases according to the respective guidelines. This study did not record any intervention that affects the olfactory function such as olfactory training and nasal irrigation either using saline or steroid. Only a few participants recorded this in a large cohort and the olfactory training was varied across the individual. Factors associated with the development of antibodies after vaccination were not assessed including the medication taken after vaccination, as some medication affects antibody formation (35). Furthermore, heterologous vaccination is not conducted in Indonesia, hence this study is unable to identify the effect of the aforementioned regimen. This study focuses on the first episode of COVID among participants. People with reinfection were excluded from the study, hence no effect of reinfection and olfactory dysfunction. The diagnosis confirmation of reinfection is challenging, particularly in identifying a true reinfection case or positive PCR results due to the virus fragment. A study addressed the association of reinfection or other terms, re-positive on disease severity and possible subsequent impacts(36). Excluding the reinfection is the limitation of this study in identifying the effect of vaccination in this specific group. The use of antiinflammation was recorded but the authors were unable to identify which kind of antiinflammation was taken by the participants. In practice, antiinflammation is not included in COVID-19 regimen, and over-the-counter purchase of this drug is not allowed. However, it is crucial to address the use of medication and its effect on olfactory dysfunction.

### Conclusion and implication to the public health measures against COVID 19

This study identified the importance of having full vaccination in preventing olfactory dysfunction if contracted with SARS Cov2 in Indonesia’s setting. However, a booster shot is recommended to those who received the full doses of inactivated viral vaccine to be protected from the olfactory dysfunction if infected. The recommended day should be more than 88 days after receiving the second dose. This booster policy of people who received inactivated vaccines has been conducted in several countries. Thailand has initiated a clear guideline to provide the booster using viral-vector vaccine 90 days after the second vaccination of inactivated virus vaccine and this could be applied to Indonesia setting or other countries who conducted the vaccination program mainly with the inactivated regimen. Redefining the full vaccination for those who receive two doses of inactivated vaccine to be at least three doses seems to be inevitable. In summary, Vaccination will reduce the burden due to olfactory dysfunction and improve the quality of life among people infected with SARS Cov2.

## Data Availability

All data produced in the present study are available upon reasonable request to the authors

## Acknowledgment

The authors would like to express gratitude to the covidsurvivor.id, the largest COVID survivor group Indonesia with its leader Juno Simorangkir, and Medical Youth Research Club Faculty of Medicine Hasanuddin University Indonesia, and all the participants who contribute to the research.

## Availability of the data

The data is available upon request

## References

1. Organization WH. WHO Coronavirus (COVID-19) Dashboard, Situation By Region, Country, Territory, and Area Geneva 2021 [Available from: https://covid19.who.int/table.

2. Lippi G, Wong J, Henry BM. Hypertension in patients with coronavirus disease 2019 (COVID-19): a pooled analysis. Polish archives of internal medicine. 2020;130(4):304–9.

3. Leung C. Clinical features of deaths in the novel coronavirus epidemic in China. Reviews in medical virology. 2020;30(3):e2103.

4. Tenforde MW, Kim SS, Lindsell CJ, Billig Rose E, Shapiro NI, Files DC, et al. Symptom Duration and Risk Factors for Delayed Return to Usual Health Among Outpatients with COVID-19 in a Multistate Health Care Systems Network - United States, March-June 2020. MMWR Morbidity and mortality weekly report. 2020;69(30):993–8.

5. Stavem K, Ghanima W, Olsen MK, Gilboe HM, Einvik G. Persistent symptoms 1.5-6 months after COVID-19 in non-hospitalised subjects: a population-based cohort study. Thorax. 2021;76(4):405–7.

6. Health NIf, 2 CEJEr, prevalence. COVID-19 Rapid Guideline: Managing the Long-Term Effects of COVID-19).(NG188). 2020.

7. Fernández-de-Las-Peñas C, Palacios-Ceña D, Gómez-Mayordomo V, Cuadrado ML, Florencio LL. Defining Post-COVID Symptoms (Post-Acute COVID, Long COVID, Persistent Post-COVID): An Integrative Classification. International journal of environmental research and public health. 2021;18(5).

8. Lopez-Leon S, Wegman-Ostrosky T, Perelman C, Sepulveda R, Rebolledo PA, Cuapio A, et al. More than 50 long-term effects of COVID-19: a systematic review and meta-analysis. Scientific Reports. 2021;11(1):16144.

9. Coelho DH, Reiter ER, Budd SG, Shin Y, Kons ZA, Costanzo RM. Quality of life and safety impact of COVID-19 associated smell and taste disturbances. American journal of otolaryngology. 2021;42(4):103001.

10. Poerbonegoro NL, Reksodiputro MH, Sari DP, Mufida T, Rahman MA, Reksodiputro LA, et al. Cross-sectional study on the proportion of smell and taste disturbances in hospitalized COVID-19 patients. Annals of Medicine and Surgery. 2021;71:102909.

11. van Riel D, Verdijk R, Kuiken T. The olfactory nerve: a shortcut for influenza and other viral diseases into the central nervous system. The Journal of pathology. 2015;235(2):277–87.

12. Hoffmann M, Kleine-Weber H, Schroeder S, Krüger N, Herrler T, Erichsen S, et al. SARS-CoV-2 Cell Entry Depends on ACE2 and TMPRSS2 and Is Blocked by a Clinically Proven Protease Inhibitor. Cell. 2020;181(2):271–80.e8.

13. Torabi A, Mohammadbagheri E, Akbari Dilmaghani N, Bayat AH, Fathi M, Vakili K, et al. Proinflammatory Cytokines in the Olfactory Mucosa Result in COVID-19 Induced Anosmia. ACS chemical neuroscience. 2020;11(13):1909–13.

14. Brann DH, Tsukahara T, Weinreb C, Lipovsek M, Van den Berge K, Gong B, et al. Non-neuronal expression of SARS-CoV-2 entry genes in the olfactory system suggests mechanisms underlying COVID-19-associated anosmia. Science advances. 2020;6(31).

15. Butowt R, Bilinska K. SARS-CoV-2: Olfaction, Brain Infection, and the Urgent Need for Clinical Samples Allowing Earlier Virus Detection. ACS chemical neuroscience. 2020;11(9):1200–3.

16. Kabbani N, Olds JL. Does COVID19 Infect the Brain? If So, Smokers Might Be at a Higher Risk. Molecular pharmacology. 2020;97(5):351–3.

17. Yuan F, Huang T, Wei Y, Wu D. Steroids and Olfactory Training for Postviral Olfactory Dysfunction: A Systematic Review. Frontiers in neuroscience. 2021;15:708510.

18. COVID 19 Situation Indonesia [press release]. Jakarta, 13 December 2021 2021.

19. Organization WH. Diagnostic testing for SARS-CoV-2. Geneva: World Health Organization; 2020 11 September 2020.

20. Organization WH. Antigen-detection in the diagnosis of SARS-CoV-2 infection. 2021.

21. Kritikos A, Caruana G, Brouillet R, Miroz JP, Abed-Maillard S, Stieger G, et al. Sensitivity of Rapid Antigen Testing and RT-PCR Performed on Nasopharyngeal Swabs versus Saliva Samples in COVID-19 Hospitalized Patients: Results of a Prospective Comparative Trial (RESTART). Microorganisms. 2021;9(9).

22. World Health O. Criteria for releasing COVID-19 patients from isolation: scientific brief, 17 June 2020. Geneva: World Health Organization; 2020 2020. Contract No.: WHO/2019-nCoV/Sci_Brief/Discharge_From_Isolation/2020.1.

23. Mathuria JP, Yadav R, Rajkumar. Laboratory diagnosis of SARS-CoV-2 - A review of current methods. Journal of infection and public health. 2020;13(7):901–5.

24. Liu Q, Qin C, Liu M, Liu J. Effectiveness and safety of SARS-CoV-2 vaccine in real-world studies: a systematic review and meta-analysis. Infectious Diseases of Poverty. 2021;10(1):132.

25. Zou LQ, Linden L, Cuevas M, Metasch ML, Welge-Lüssen A, Hähner A, et al. Self-reported mini olfactory questionnaire (Self-MOQ): A simple and useful measurement for the screening of olfactory dysfunction. The Laryngoscope. 2020;130(12):E786–e90.

26. COVID-19 Distribution according to provinces [Internet]. 2021 [cited 15 December 2021]. Available from: https://covid19.go.id/peta-sebaran.

27. Leso V, Fontana L, Iavicoli I. Susceptibility to Coronavirus (COVID-19) in Occupational Settings: The Complex Interplay between Individual and Workplace Factors. International journal of environmental research and public health. 2021;18(3).

28. Bagnasco D, Passalacqua G, Braido F, Tagliabue E, Cosini F, Filauro M, et al. Quick Olfactory Sniffin’ Sticks Test (Q-Sticks) for the detection of smell disorders in COVID-19 patients. The World Allergy Organization journal. 2021;14(1):100497.

29. Indonesia MoH. Genome Sequencing Results of SARS COV2 January-September 2021. In: Health P, editor. Jakarta 2021.

30. Ong SWX, Chiew CJ, Ang LW, Mak T-M, Cui L, Toh MPHS, et al. Clinical and Virological Features of Severe Acute Respiratory Syndrome Coronavirus 2 (SARS-CoV-2) Variants of Concern: A Retrospective Cohort Study Comparing B.1.1.7 (Alpha), B.1.351 (Beta), and B.1.617.2 (Delta). Clinical Infectious Diseases. 2021.

31. Cohn BA, Cirillo PM, Murphy CC, Krigbaum NY, Wallace AW. SARS-CoV-2 vaccine protection and deaths among US veterans during 2021.0(0):eabm0620.

32. Abdelmaksoud AA, Ghweil AA, Hassan MH, Rashad A, Khodeary A, Aref ZF, et al. Olfactory Disturbances as Presenting Manifestation Among Egyptian Patients with COVID-19: Possible Role of Zinc. Biological trace element research. 2021;199(11):4101–8.

33. Parnham MJ, Haber VE, Giamarellos-Bourboulis EJ, Perletti G, Verleden GM, Vos R. Azithromycin: Mechanisms of action and their relevance for clinical applications. Pharmacology and Therapeutics. 2014;143(2):225–45.

34. Li X, Zhong X, Wang Y, Zeng X, Luo T, Liu Q. Clinical determinants of the severity of COVID-19: A systematic review and meta-analysis. PLOS ONE. 2021;16(5):e0250602.

35. Bancos S, Bernard MP, Topham DJ, Phipps RP. Ibuprofen and other widely used non-steroidal anti-inflammatory drugs inhibit antibody production in human cells. Cellular immunology. 2009;258(1):18–28.

36. Slezak J, Bruxvoort K, Fischer H, Broder B, Ackerson B, Tartof S. Rate and severity of suspected SARS-Cov-2 reinfection in a cohort of PCR-positive COVID-19 patients. Clinical Microbiology and Infection. 2021;27(12):1860.e7-.e10.

